# *KRAS* mutations impact clinical outcome in metastatic non-small cell lung cancer

**DOI:** 10.1101/2021.11.27.21266822

**Authors:** Ella A. Eklund, Clotilde Wiel, Henrik Fagman, Levent M. Akyürek, Sukanya Raghavan, Jan Nyman, Andreas Hallqvist, Volkan I. Sayin

## Abstract

**Purpose:** There is an urgent need to identify new predictive biomarkers for treatment response to both platinum doublet chemotherapy (PD) and immune checkpoint blockade (ICB) with pembrolizumab. Here we evaluated whether treatment outcome could be affected by *KRAS* mutational status in patients with metastatic (stage IV) non-small cell lung cancer (NSCLC).

**Methods:** All consecutive patients molecularly assessed and diagnosed between 2016-2018 with stage IV NSCLC in the region of West Sweden were included in this multi-center retrospective study. Primary study outcome was overall survival (OS).

**Results:** Out of 580 stage IV NSCLC patients, 35.5% harbored an activating mutation in the *KRAS* gene (KRAS^MUT^). Compared to *KRAS* wild-type (KRAS^WT^), KRAS^MUT^ was a negative factor for OS (*p* = 0.014). On multivariate analysis, KRAS^MUT^ persisted as a negative factor for OS (HR 1.288, 95% CI 1.091-1.521, *p* = 0.003). When treated with first-line platinum doublet (*n* = 195), KRAS^MUT^ is a negative factor for survival (*p* = 0.018) with median OS 9 months vs KRAS^WT^ 11 months. On multivariate analysis, KRAS^MUT^ persisted as a negative factor for OS (HR 1.564, 95%CI 1.124-2.177, *p* = 0.008). KRAS^MUT^ patients with high PD-L1 expression (PD-L1^high^) had better OS than PD-L1^high^ KRAS^WT^ patients (*p* = 0.036). In response to first-line ICB, KRAS^MUT^ patients had a significant (*p* = 0.006) better outcome than KRAS^WT^ with a median OS 23 vs 6 months. On multivariable Cox analysis, KRAS^MUT^ status was an independent prognostic factor for better OS (HR 0.349, 95%CI 0.148-0.822, *p* = 0.016).

**Conclusions:** *KRAS* mutations is a positive predictive factor for treatment with pembrolizumab and a negative predictive factor for platinum doublet chemotherapy as well as general OS in stage IV NSCLC.

## Background

### Non-small cell lung cancer

(NSCLC) is the most commonly diagnosed cancer worldwide with 2.1 million new cases and 1.8 million deaths annually [1]. NSCLC can be treated effectively with local management of the primary tumor in early stages but the 5-year survival for patients with advanced metastatic (stage IV) NSCLC is below 10% [2, 3] NSCLC generally tends to have a high overall mutational burden mainly due to exposure of exogenous mutagens like tobacco smoke but also air pollution, causing high genomic instability and inter-patient heterogeneity [4].

The treatment options for stage IV NSCLC patients have become dependent on molecular profiling because of the introduction of small molecular kinase inhibitors (SMKIs) targeting activating mutations in EGFR, ALK, BRAF, RET and ROS1 oncogenes [5]. Unfortunately, the majority of stage IV patients do not harbor a targetable driving mutation, for which platinum-based chemotherapy doublets (PD) has been the only available treatment option until recently. The introduction of immune checkpoint blockade (ICB) targeting programmed death ligand 1 (PD-L1) or programmed cell death 1 (PD-1) proteins led to impressive clinical results [6-8]. Consequently, all stage IV patients without any clinically actionable mutations are assessed for ICB, single or in combination with chemotherapy, as a first-line treatment option [9-12]. PD-L1 expression is the only validated predictive marker for response to immunotherapy. Yet, its accuracy as an individual prediction tool is not as good as initially assumed since on one hand PD-L1 negative patients have been reported as ICB responders while on the other, many patients with tumors expressing high levels of PD-L1 are being reported as non-responders to ICB therapy [9, 13]. Along these lines, a meta-analysis of six randomized controlled trials with ICB showed that PD-L1 expression levels was neither prognostic nor predictive for OS [14].

The most frequent oncogenic driver in NSCLC is the Kirsten rat sarcoma viral oncogene (*KRAS*) present in up to 40% of all cases and the most common mutations are G12C, G12V and G12D [15, 16]. *KRAS* mutations have been considered to negatively influence the prognosis of NSCLC. Accordingly, *KRAS* mutations have been associated with a shorter OS following first line PD treatment in more recent studies [17-21]. Attempts to target mutant KRAS and its downstream mediators in cancer therapy remain largely unsuccessful [4, 22]. However, two inhibitors efficiently targeting mutant KRAS-G12C have been developed but the clinical efficacy is yet to be determined in ongoing trials [23-27]. Concurrently, it has been suggested that NSCLC harboring *KRAS* mutations might benefit from ICB therapy compared to KRAS wild type tumors [28]. Nevertheless, retrospective real-world data have so far shown inconclusive results [28-33].

The gradual introduction of ICB treatment in recent years dawned a new era with high hopes for patients with stage IV NSCLC. Clinical trials have shown ICB as being superior to chemotherapy in patients harboring tumors with high tumor proportion score (TPS) ≥50% for PD-L1. These results were confirmed in a recent network meta – analysis [7, 8, 10, 34]. However, the impact of mutant KRAS was not addressed in these analyses. In addition, multiple real-world studies have reported conflicting results regarding the impact of mutant KRAS on ICB response. In early days of immunotherapy, only PD-L1 ≥ 50% patients were included in first-line ICB-treatment. We know now that PD-L1 negative patients may also respond to ICB [9, 13]. Interestingly, some studies suggest that PD-L1 expression per se might have a prognostic impact on NSCLC [35, 36].

By including all consecutive patients molecularly assessed and diagnosed with stage IV NSCLC between 2016-2018 in west Sweden, the current retrospective cohort study provides a unique real-world dataset for assessing the impact of *KRAS* mutations and PD-L1 expression on OS following first-line standard of care, including platinum doublet chemotherapy and immunotherapy with pembrolizumab.

## Materials and Methods

### Patient population

We conducted a multi-centers retrospective study including all consecutive NSCLC patients diagnosed with stage IV NSCLC and having molecular assessment performed between 2016-2018 in the Region Västra Götaland (region of West Sweden), Sweden (*n* = 580). Approval from the Swedish Ethical Review Authority (Dnr 2019-04771) was obtained prior to study commencement. All patients with available tissue sample where an adenocarcinoma component could not be excluded were systematically assessed with NGS for known genetic drivers (see below) within clinical praxis. Patient demographics (including age, gender, Eastern Cooperative Oncology Group (ECOG) performance status and smoking history), cancer stage, number of metastasis locations, pathological details (histology, mutation status including *KRAS* mutational status and subtype), treatment and outcome data were retrospectively collected from patient charts and the Swedish Lung Cancer Registry.

### Mutational status

Patients were assessed with NGS for mutational status on DNA from FFPE blocks or cytological smears using the Ion AmpliSeq™ Colon and Lung Cancer Panel v2 from Thermo Fisher Scientific as a part of the diagnostic workup process at the Department of Clinical Pathology at Sahlgrenska University Hospital, assessing hotspot mutations in EGFR, BRAF, KRAS and NRAS. Until June 2017, ALK-fusions were assessed with immunohistochemistry (IHC), and with fluorescence in situ hybridization (FISH) if positive or inconclusive IHC; ROS1 was analyzed upon request with FISH. Thereafter, ALK, ROS1 and RET fusions were assessed on RNA using the Oncomine Solid Tumor Fusion Panel from Thermo Fisher Scientific.

### PD-L1 expression

Programmed death ligand 1 (PD-L1) expression was determined based on percentage of tumor cells with positive membranous staining and was reported as the tumor proportion score (TPS). PD-L1 negative TPS <1%, low TPS 1%-49%, high TPS ≥50%. PD-L1 expression was detected using the PD-L1 IHC 28-8 pharmDx system during routine diagnostic workup and staining was assessed by lung pathologists.

### ICB treatment

During the time period of this study, the only ICB treatment approved for first-line treatment was Pembrolizumab, a humanized antibody targeting PD-1. The criteria was PD-L1^high^ TPS ≥50% for first-line and PD-L1^low^ TPS ≥1% for second line treatment.

### PD treatment

Platinum doublet treatment consists of carboplatin or cisplatin in combination with one more non-platinum chemotherapy agent such as pemetrexed, vinorelbin, gemcitabine, paclitaxel, etoposide or vincristine.

### Study objectives

The primary outcome of this study was OS, defined as the interval between the date of diagnostic sample collection and the date of death from any cause. Patients alive or lost to follow-up at data were censored at last contact. Median follow up time was 7 months.

We compared OS stratified on KRAS^WT^ and KRAS^MUT^ for the entire cohort, for all patients receiving life extending treatment excluding patients receiving best supportive care or palliative radiotherapy (e.g., single tumor radiation for pain relief), and for all patients receiving PD or ICB as first-line treatment. We also investigated the impact of TPS score (negative, low and high) on OS, stratified on KRAS mutational status.

### Statistical analysis

Clinical characteristics were summarized using descriptive statistics and evaluated with univariate analysis. Kaplan Meier survival curves were generated to assess OS. Log-rank test was used to assess significant differences in OS between groups. Multivariable Cox regression analysis was conducted to compensate for potential confounders. Statistical significance was set at *p*<0.05 and no adjustments were made for multiple comparisons. One-sided Fisher’s Exact Test was used to determine the connection between KRAS status to being alive in the ICB-treated group. Data analysis was conducted using IBM SPSS Statistics version 27 and GraphPad Prism version 9.

## Results

### Patients and tumor characteristics

A total of 597 consecutive patients diagnosed with stage IV NSCLC were molecularly assessed during 2016-2018 in West Sweden. A total of 17 patients were excluded for having incomplete medical records, for receiving palliative treatment before 2016, for incorrect diagnosis or for receiving simultaneous treatment for another type of cancer (Figure. 1A). Among the 580 included patients, more than a third harbored a *KRAS* mutation (206, 35.5%), the majority were female (326, 56.2%), the median age was 71 years and 80 % were current or former smokers (Table 1). The majority of patients (322, 55.5%) had a good Performance Status (PS) with low ECOG 0-1 at diagnosis (Table 1). Histologically, the vast majority of NSCLCs were adenocarcinoma of the lung (498, 85.9%) while squamous cell carcinoma cases were relatively low which was expected due to the selection of histological type for NGS assessment (32, 5.5%) (Table 1). In line with earlier studies, patients diagnosed with a low ECOG score had a significantly better overall survival (OS), compared to patients diagnosed with a higher ECOG score (*p* = 0.0001) (Supplemental Figure. 1A) [3, 37]. In addition, higher number of metastasis locations at diagnosis correlated with poor OS (*p* = 0.0002) (Supplemental Figure. 1B), which was expected [3, 37]. Furthermore, there was a trend towards better OS for females compared to males (*p* = 0.064) (Supplemental Figure. 1C) [37, 38]. When comparing the baseline characteristics of KRAS^WT^ with KRAS^MUT^ patients, there were more females and a higher proportion of current and former smokers in the KRAS^MUT^ population (Table 1).

**Figure 1.**
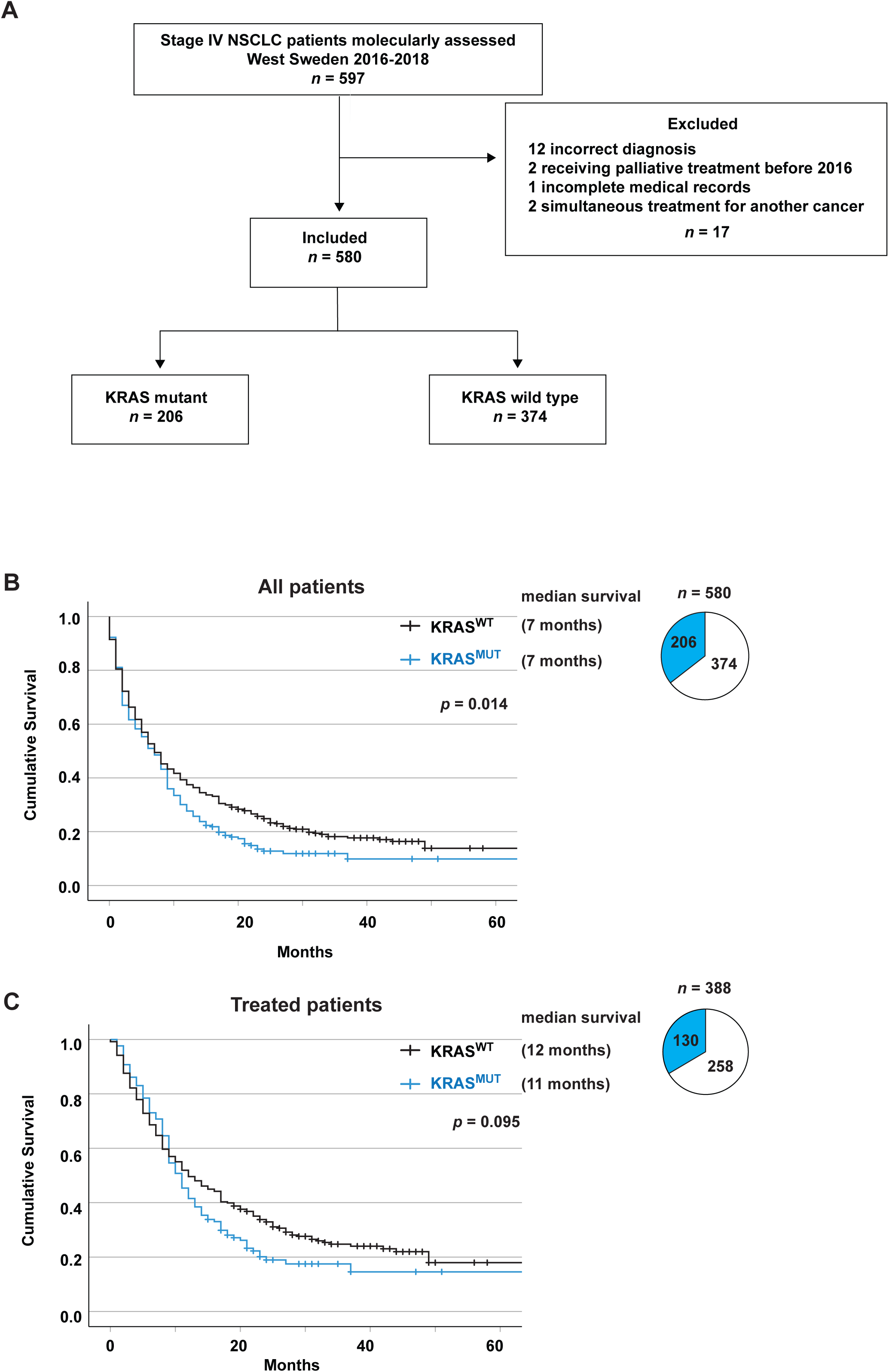
*KRAS* mutations is a negative factor for overall survival. A. Flow chart showing the patient selection for the study. B. Kaplan-Meier estimates comparing overall survival and median survival stratified on KRAS^WT^ vs KRAS^MUT^ for the full cohort (*n* = 580). Pie chart showing patient distribution between KRAS^WT^ and KRAS^MUT^. C. Kaplan-Meier estimates comparing overall survival for all receiving life extending treatment (no treatment and only palliative radiotherapy excluded) stratified on KRAS^WT^ vs KRAS^MUT^. Pie chart showing patient distribution between KRAS^WT^ and KRAS^MUT^.

### *KRAS* mutations is a negative factor for overall survival in stage IV NSCLC

Survival estimates for the whole study cohort displayed that *KRAS* mutations was a significant negative factor for overall survival (*p* = 0.014) (Figure. 1B). On multivariate Cox regression analysis, KRAS^MUT^ status and male sex were independent prognostic factors for worse OS (HR 1.288, 95%CI 1.091-1.521, *p* = 0.003 and HR 1.179, 95%CI 1.009-1.378, *p* = 0.038). The population that received any type of life extending treatment (*n* = 388), excluding patients only receiving best supportive care or palliative radiotherapy (e.g., single tumor radiation for pain relief) did not show a significant difference in OS between KRAS^MUT^ and KRAS^WT^ (*p* = 0.095) (Figure. 1C).

When treated with first-line PD, one of standard treatments at the time, (*n* = 219), KRAS^MUT^ is a significant negative factor for survival (*p* = 0.001) with median OS 9 months vs KRAS^WT^ 14 months (Figure. 2A). Baseline characteristics of the patients show that around 20% of KRAS^WT^ patients displayed a druggable genomic alterations in ALK or EGFR (Supplemental Table 1). To eliminate the risk of these patients driving any difference between KRAS^MUT^ and KRAS^WT^ groups due to potentially receiving targeted therapies in the second line of treatment, we excluded all patients with EGFR mutations and ALK fusions from the KRAS^WT^ group from further analysis. Nevertheless, in patients receiving first-line PD chemotherapy (*n* = 195), KRAS^MUT^ is a significant (*p* = 0.018) negative factor for survival (Figure. 2B), with median OS 9 months for KRAS^MUT^ vs 11 months for KRAS^WT^. In multivariate Cox regression analysis, KRAS^MUT^ status and ECOG 2 or higher were independent prognostic factors for worse OS (HR 1.564, 95%CI 1.124-2.177, *p* = 0.008 and HR 1.906, 95%CI 1.307-7,779, *p* <0.001).

**Figure 2.**
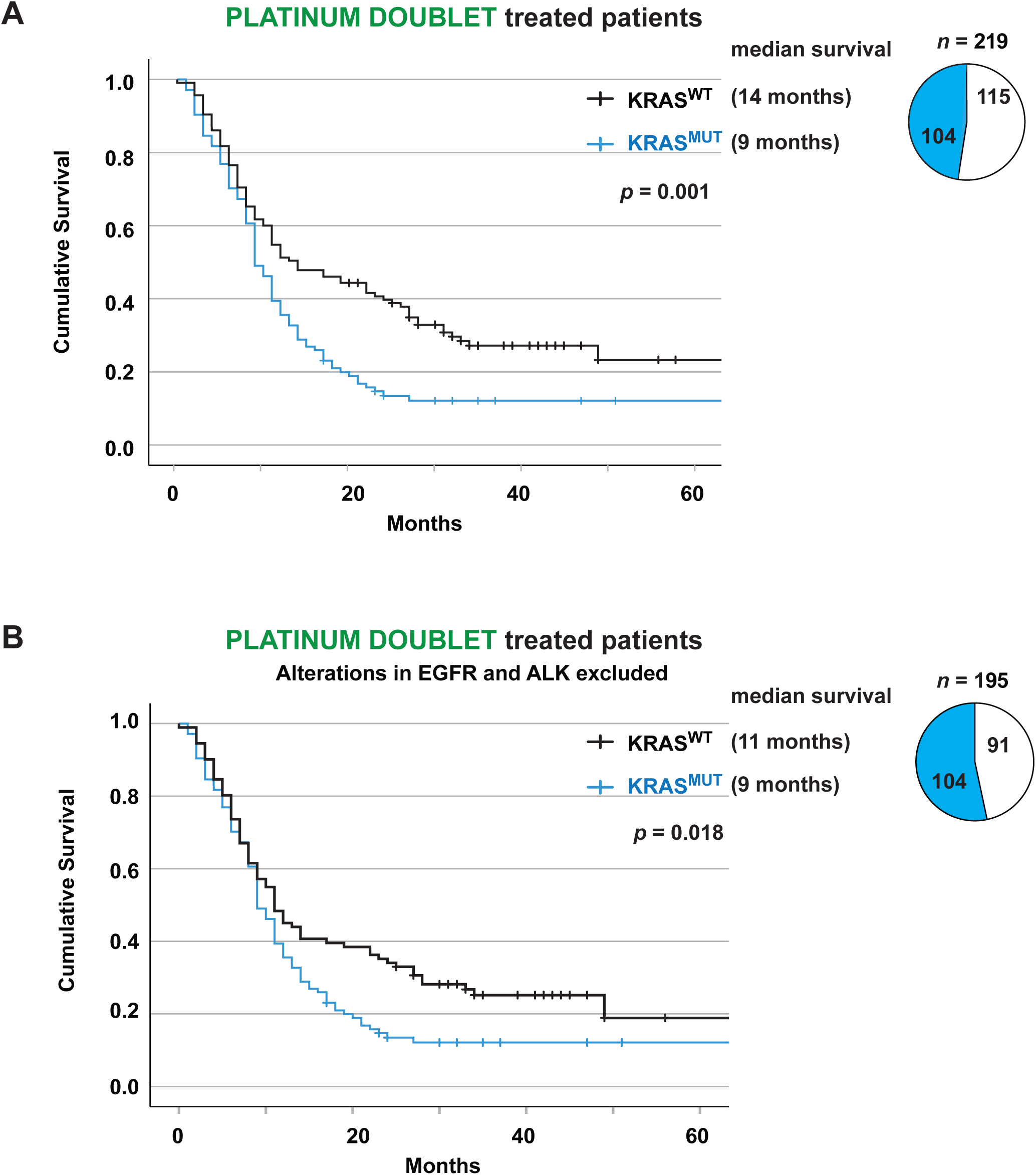
KRAS^MUT^ is a negative factor for overall survival when treated with platinum doublet independently of EGFR and ALK mutations. A. Kaplan-Meier estimates comparing overall survival for all receiving treatment with platinum doublet stratified on KRAS^WT^ vs KRAS^MUT^. Pie chart showing patient distribution between KRAS^WT^ and KRAS^MUT^. B. Kaplan-Meier estimates comparing overall survival for all receiving treatment with platinum doublet stratified on KRAS^WT^ vs KRAS^MUT^. Patients with EGFR and ALK alterations are excluded from KRAS^WT^ group. Pie chart showing patient distribution between KRAS^WT^ and KRAS^MUT^.

### Overall survival for stage IV NSCLC treated with first-line PD or ICB stratifies based on *KRAS* mutational status

When comparing OS for all stage IV patients receiving PD (*n* =195) or ICB (*n* = 37) in a first-line setting, there was no difference between the two treatment groups (*p* = 0.678), median OS 11 months for PD vs 13 months for ICB (Figure. 3A). KRAS^MUT^ patients treated with ICB (*n* = 20) had a significantly (*p* = 0.003) better outcome compared to patients treated with PD (*n* = 104), median OS 23 vs 9 months (Figure. 3B). KRAS^WT^ patients treated with ICB (*n* = 17) had significantly (*p* = 0.023) worse survival than patients treated with PD (*n* = 91,) median OS 6 vs 11 months (Figure. 3C).

**Figure 3.**
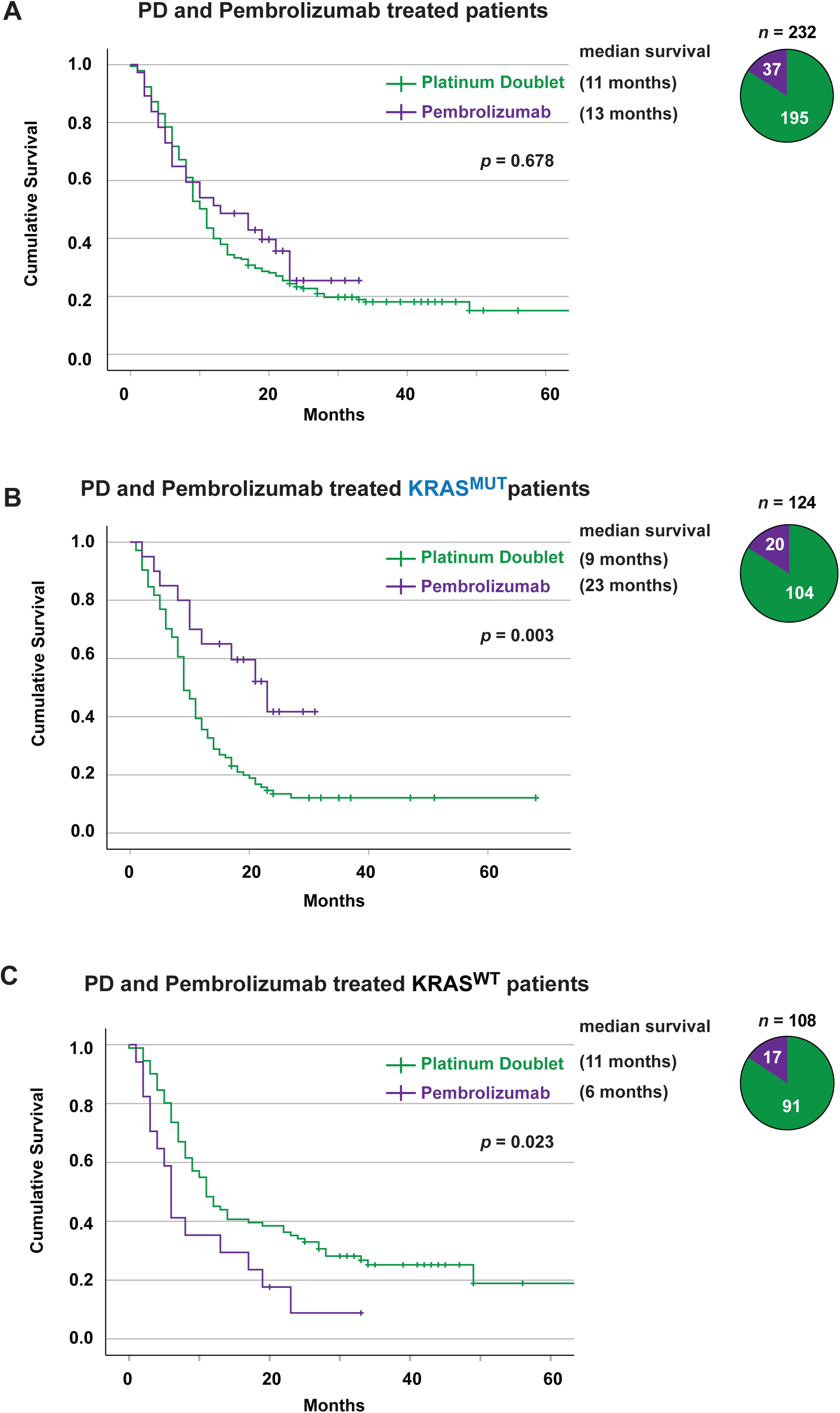
KRAS^MUT^, but not KRAS^WT^, has a better outcome on ICB than platinum doublet treatment. A. Kaplan-Meier estimates comparing overall survival for all patients, excluding patients with ALK or EGFR alterations, receiving treatment with PD vs ICB. Pie chart showing patient distribution between PD and ICB. B. Kaplan-Meier estimates comparing overall survival for all KRAS^MUT^ receiving treatment with PD vs ICB. Pie chart showing patient distribution between platinum doublet and ICB. C. Kaplan-Meier estimates comparing overall survival for KRAS^WT^, excluding patients with ALK or EGFR alterations, receiving treatment with platinum doublet vs ICB. Pie chart showing patient distribution between PD and ICB. ICB: Immune Checkpoint Blockade; PD: Platinum Doublet

### PD-L1 expression in tumors from stage IV patients is a positive factor for overall survival in KRAS^MUT^ but not KRAS^WT^ patients

Currently, tumoral PD-L1 expression is the only biomarker for ICB treatment widely used in the clinic for NSCLC [39, 40]. When analyzing all treated stage IV NSCLC patients with known PD-L1 status (*n* = 261) stratified on *KRAS* mutational status and PD-L1 expression levels (negative, low or high) (Figure. 4A), we observed that increased PD-L1 expression correlated with significant better OS in KRAS^MUT^ patients (*p* = 0.036, Figure 4B). KRAS^MUT^ patients clearly separate on OS between PD-L1 negative, low and high groups with median OS 6, 11 and 17 months respectively (Figure. 4B). No correlation between PD-L1 expression and OS in KRAS^WT^ patients was observed (Figure. 4C). There was a larger proportion of PD-L1^high^ in the KRAS^MUT^ population compared to KRAS^WT^ (43.0% vs 32.7%).

**Figure 4.**
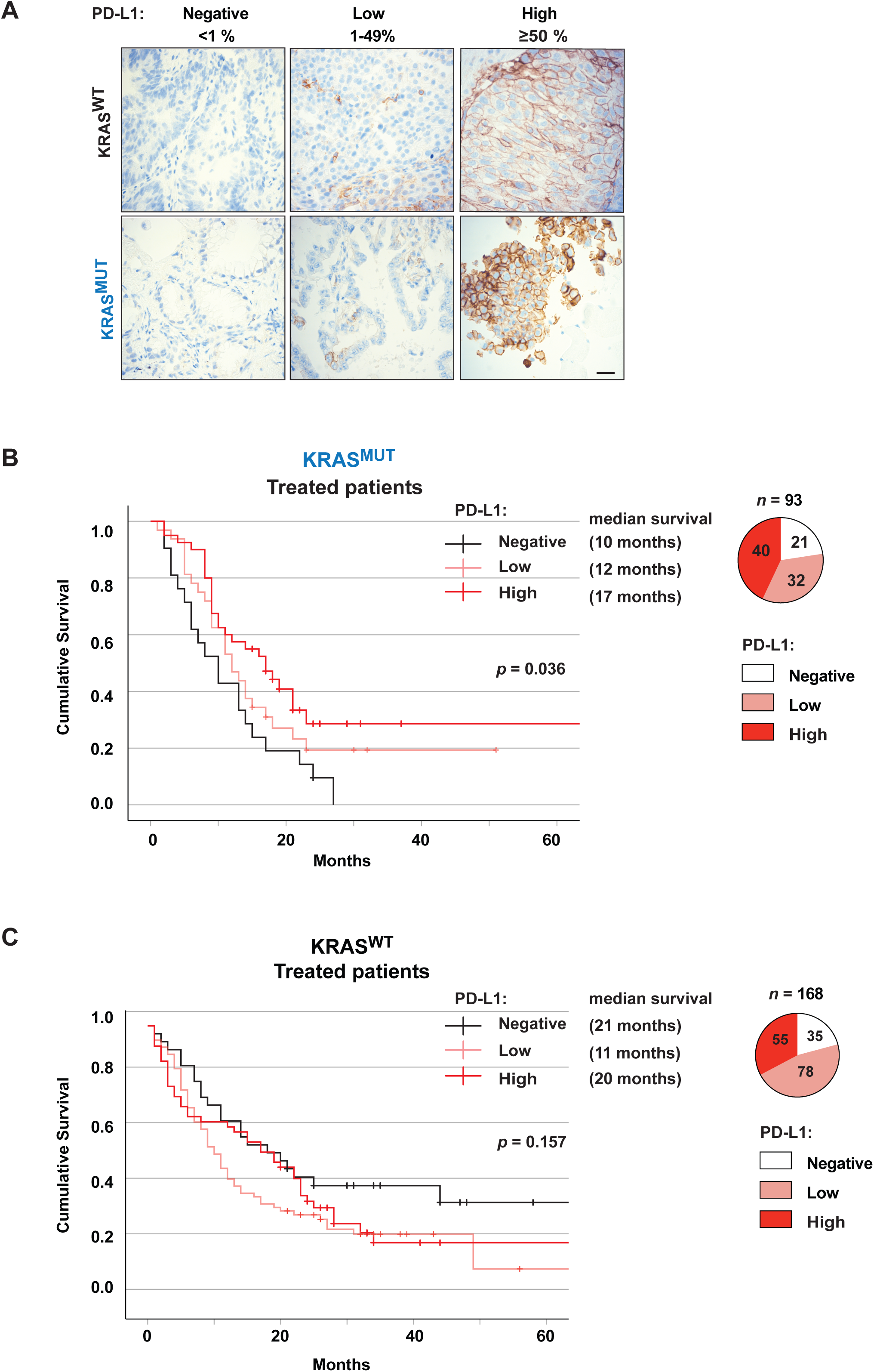
PD-L1 status has an impact on overall survival for treated patients with KRAS^MUT^. A. IHC depicting examples of PD-L1 staining used to assess PD-L1 status in both KRAS^WT^ and KRAS^MUT^ NSCLC patients. PD-L1 expression is classified as negative, < 50% or ≥50% based on tumor proportion score. B. Kaplan-Meier estimates comparing overall survival for KRAS^MUT^ treated patients stratified on PD-L1 status. Pie chart showing patient distribution between PD-L1 negative, <50% and ≥50%. C. Kaplan-Meier estimates comparing overall survival for KRAS^WT^ treated patients stratified on PD-L1 status. Pie chart showing patient distribution between PD-L1 negative, <50% and ≥50%. IHC: Immunohistochemistry; PD-L1: Programmed Death Ligand 1

### *KRAS* mutations are a positive factor for overall survival in patients with PD-L1^high^ tumors receiving immunotherapy

We next analyzed the specific impact of PD-L1^high^ expression on OS comparing all PD-treated vs ICB-treated patients: outcome was significantly improved for KRAS^MUT^ patients on ICB treatment (*p* = 0.028) with median OS being 9 vs 23 months (Figure 5A). KRAS^WT^ PD-L1^high^ patients displayed a worse outcome for patients on ICB treatment compared to PD treatment (*p* = 0.010) with median OS 6 months vs 28 months for each respective treatment (Figure. 5B). Importantly, the KRAS^MUT^ group had a significantly (*p* = 0.006) better outcome on ICB treatment compared to the KRAS^WT^ group with a median OS 23 vs 6 months (Figure. 5C, Supplemental Table 2). On multivariate Cox regression analysis, KRAS^MUT^ status and ECOG PS 0-1 were independent prognostic factors for better OS following first-line ICB (HR 0.349, 95%CI 0.148-0.822, *p* = 0.016 and HR 0.398, 95%CI 0.165-0.963, *p* = 0.041). Finally, when looking at all patients treated with pembrolizumab individually, there were significantly more KRAS^MUT^ patients alive at last follow-up compared to KRAS^WT^ patients (50% vs 11.8%, *p* = 0.0152) (Figure 5D).

**Figure 5.**
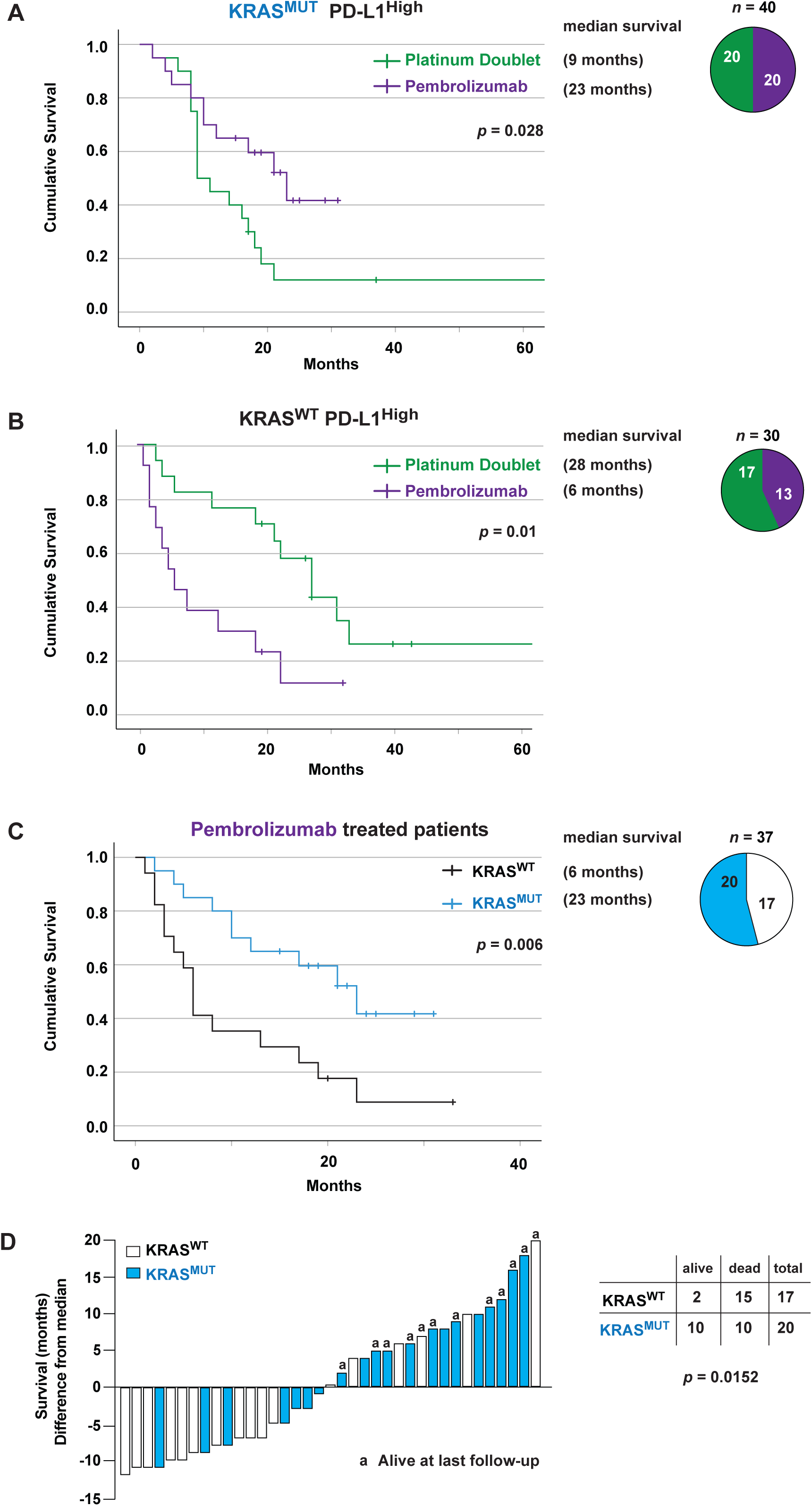
KRAS^MUT^ patients have a better outcome on ICB treatment than KRAS^WT^ patients. A. Kaplan-Meier estimates comparing overall survival for KRAS^MUT^ PD-L1 ≥50% treated patients receiving PD or ICB. Pie chart showing patient distribution receiving PD or ICB treatment. B.Kaplan-Meier estimates comparing overall survival for KRAS^WT^ PD-L1≥50% treated patients receiving PD or ICB. Pie chart showing patient distribution receiving PD or ICB treatment. C. Kaplan-Meier estimates comparing overall survival for all patients receiving ICB treatment in a first-line setting. Pie chart showing patient distribution between KRAS^WT^ and KRAS^MUT^. D. Waterfall plot showing survival time (difference from median survival) for both KRAS^WT^ and KRAS^MUT^ pembrolizumab-treated patients. Status alive at last follow-up is shown. ICB: Immune Checkpoint Blockade; PD: Platinum Doublet; PD-L1: Programmed Death Ligand 1

## Discussion

The predictive value of *KRAS* mutations for survival in stage IV NSCLC patients is still not clearly defined. While some studies have reported *KRAS* mutations to be a negative factor for survival estimates [3, 22, 41], others have shown this not to be the case [42]. The current retrospective cohort study shows a clear difference in OS for stage IV NSCLC patients when stratified on *KRAS* mutational status. Previous studies have shown that *KRAS* mutations were associated with a shorter OS in response to chemotherapy [18, 19]. In line with these studies, our data show that KRAS^MUT^ is a potential predictive negative factor for PD treatment. This is of importance, especially in light of todays’ new treatment options available such as immunotherapy. Indeed, when stratifying stage IV NSCLC patients receiving first line pembrolizumab monotherapy on *KRAS* mutational status, there is a clear increase in survival for KRAS^MUT^ patients.

Translational studies have suggested that KRAS^MUT^ patients might respond well to ICB due to a larger proportion of smokers and a higher immunologically active tumor environment as a consequence of constitutive activation of KRAS and downstream signaling pathways [28-32, 37, 43, 44]. In agreement, our data shows that first-line pembrolizumab monotherapy led to a clinically meaningful improvement in OS for KRAS^MUT^ patients when compared to first-line platinum doublet treatment. Furthermore, PD-L1 status had a clear and significant impact on stratifying OS in the KRAS^MUT^ group receiving treatment whereas no difference was observed for the KRAS^WT^ group, which is in line with a previous study reporting a similar trend for KRAS^MUT^ NSCLC receiving ICB treatment [33].

Interestingly, KRAS^WT^ patients showed a better response to PD treatment than pembrolizumab monotherapy. This might be explained by KRAS^WT^ being a heterogeneous group with the majority of patients not having an identified driving mutation, especially as patients with targetable alterations in ALK and EGFR were excluded from further analysis.

There are several retrospective studies showing different outcomes when comparing KRAS^MUT^ and KRAS^WT^ response to pembrolizumab. These discrepancies could be explained by major study design differences such as inclusion of several different stages at diagnosis, multiple treatment lines considered and inclusion of patients with no known PD-L1 status [28-33, 37]. A recent registry study from the Netherlands reported no difference in OS for PD-L1^high^ patients receiving first-line pembrolizumab monotherapy but the fraction of patients where *KRAS* mutations status had been assessed and reported to the registry was uncertain and could only be approximated (at around 75%) [37]. However, a meta-analysis of randomized controlled trials showed a significant OS benefit for KRAS^MUT^ patients in first-line immunotherapy with or without chemotherapy vs. chemotherapy alone [44], which is in line with the current study. Among these, the retrospective analysis of the phase 3 study KEYNOTE042 indicated a clear trend towards better OS in the KRAS^MUT^ group when comparing first-line pembrolizumab versus platinum-containing chemotherapy (28 vs 11 months) than for the patients with KRAS^WT^ (15 vs 12 months) [43], albeit only 301 out of 782 patients were molecularly assessed for KRAS status among which 69 KRAS^MUT^ cases were identified. Importantly, KEYNOTE042 did not address the impact of *KRAS* mutations on PD treatment outcome. In our cohort, 580 patients were assessed and 206 KRAS^MUT^ patients were identified. To the best of our knowledge, our retrospective cohort study is the largest to date comparing response to first-line ICB and PD therapy in stage IV NSCLC patients where the *KRAS* mutational status is known for all patients. Currently, pembrolizumab is a first-line immunotherapy option for all NSCLC patients with PD-L1^high^ expression independently of *KRAS* mutational status. Our study suggests that additional biomarkers for ICB response beyond PD-L1^high^ is clearly warranted, in line with other studies [9, 13, 14]. Indeed, KRAS^WT^ patients with PD-L1^high^ tumoral expression responded better to PD chemotherapy than to pembrolizumab. In contrast, KRAS^MUT^ patients clearly benefited more from pembrolizumab treatment (50% alive at last follow-up). Our results suggest that tumoral PD-L1^high^ expression in combination with *KRAS* mutations is a better predictive biomarker for ICB with pembrolizumab in stage IV NSCLC than PD-L1^high^ expression alone.

## Limitations

Although this study provides real-world evidence of the impact of *KRAS* mutational status on first-line therapy in a large and well controlled group of stage IV NSCLC patients, it is obviously limited by its retrospective nature. Despite the fact that all stage IV NSCLC patients molecularly assessed in the region of West Sweden 2016-2018 were included in this cohort study, a relatively small number of patients were treated with first-line ICB therapy when compared to first-line PD therapy. Hence pooled analyses from multi-cohort studies will be important to validate and expand our findings.

## Conclusion

Here we report that *KRAS* mutations are a positive predictive factor for immunotherapy with pembrolizumab and a negative predictive factor for platinum doublet chemotherapy as well as general OS in stage IV NSCLC. Our findings suggest that *KRAS* mutations combined with PD-L1^high^ expression is a better predictive biomarker than only PD-L1^high^ for response to first-line immunotherapy with pembrolizumab in patients with stage IV NSCLC.

## Data Availability

All data produced in the present study are available upon reasonable request to the authors

## List of abbreviations

ECOG: Eastern Cooperative Oncology Group;
ICB: Immune Checkpoint Blockade;
NSCLC: Non-Small Cell Lung Cancer;
NGS: Next Generation Sequencing;
PD: Platinum Doublet;
PD-1: Programmed Cell Death 1;
PD-L1: Programmed Death Ligand 1;
PS: Performance Status;
OS: Overall Survival;
TPS: Tumor Proportion Score

## Acknowledgements

We thank Sayin lab members for critical reading of the manuscript. In addition, we thank members of the of the Swedish Lung Cancer Registry, and the continuous reporting by Swedish healthcare employees.

## Figure legends

**Table 1** Characteristics of the total cohort as well as stratified on KRAS^WT^ and KRAS^MUT^. Data are presented as n (%).

ECOG PS, Eastern Cooperative Oncology Group Performance Status.

**Supplemental Figure 1**

**A.** Kaplan-Meier estimates comparing overall survival for the full cohort stratified on ECOG PS. Pie chart showing patient distribution between ECOG stages.

**B.** Kaplan-Meier estimates comparing overall survival for the full cohort stratified on number of metastasis locations. Pie chart showing patient distribution between number of metastasis locations.

**C.** Kaplan-Meier estimates comparing overall survival for the full cohort stratified on gender. Pie chart showing patient distribution between male and female.

ECOG: Eastern Cooperative Oncology Group; PS: Performance Status

**Supplemental Table 1**

(Related to Figure. 2A) Characteristics of all PD treated patients.

**Supplemental Table 2**

(Related to Figure 5A) Characteristics of all patients treated with first-line ICB

