## Supplementary figures and images for "*KRAS* mutations impact clinical outcome in metastatic non-small cell lung cancer"

### Supplemental Figure 1

#### Eklund et al. Supplemental Figure 1

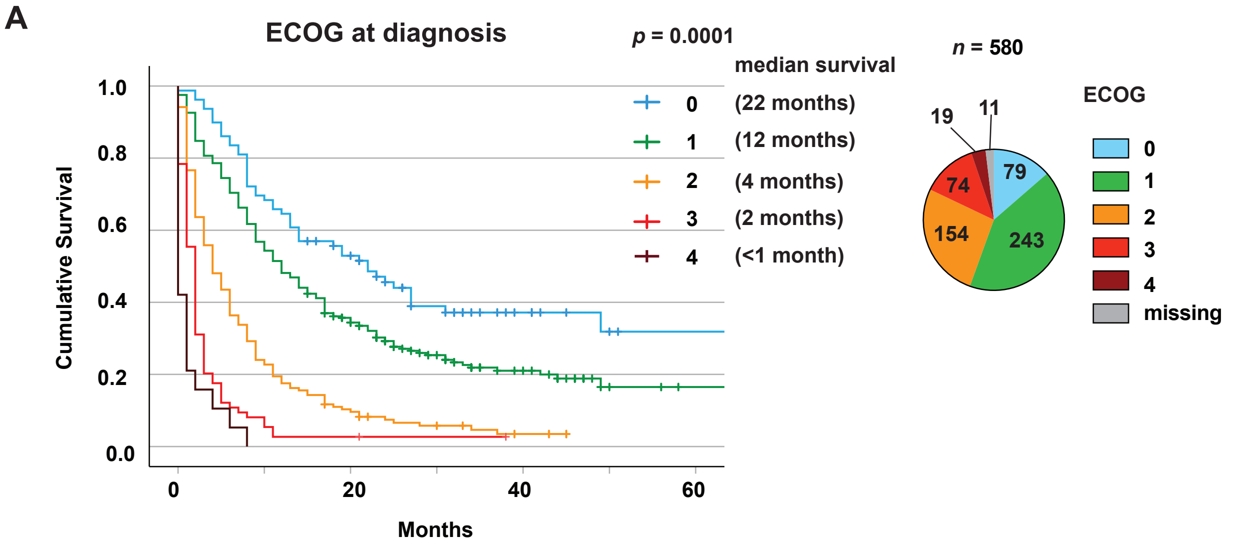


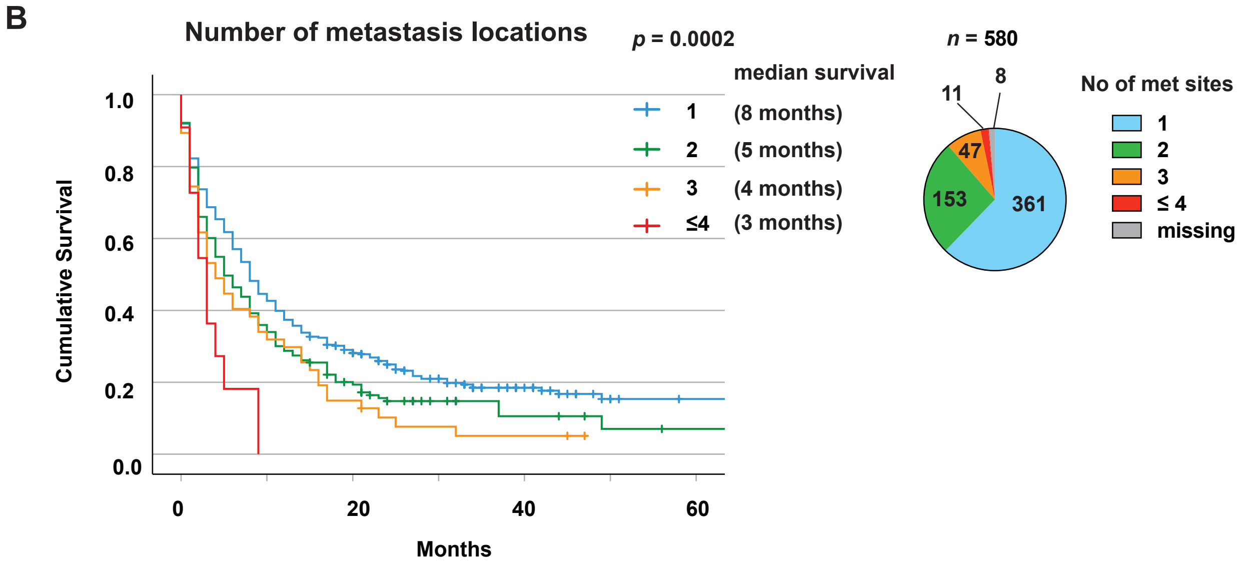


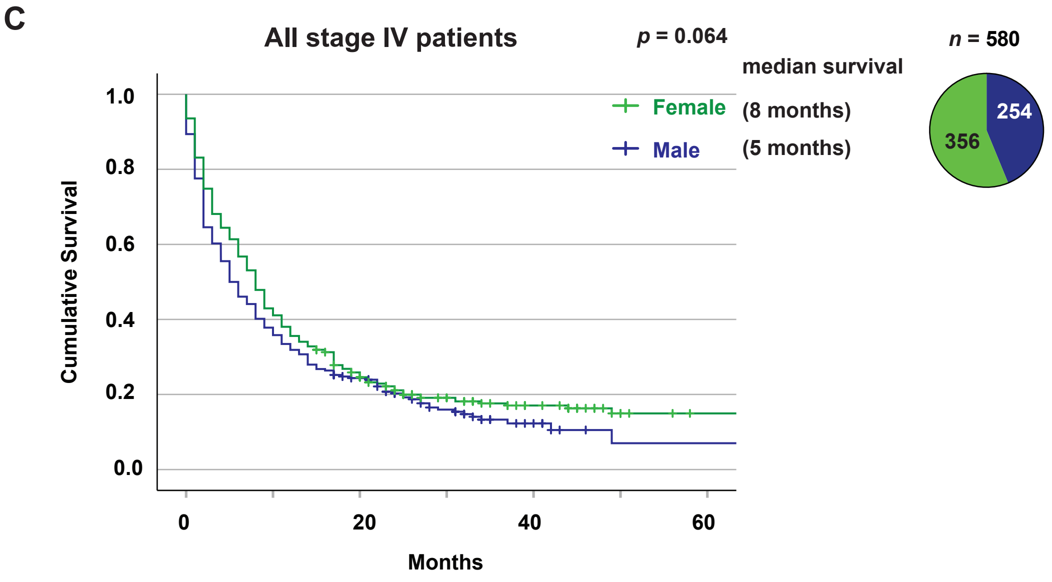
